# The development and usability testing of two arts-based knowledge translation tools for pediatric asthma

**DOI:** 10.1101/2023.02.23.23286192

**Authors:** Shannon D. Scott, Lisa Hartling

## Abstract

Asthma is the most common chronic condition in children with an estimated 15% of children and youth living with asthma in Canada. Acute asthma exacerbations, or asthma attacks, are the main reason for children to seek emergency care, contributing to financial burdens for families and healthcare systems. This burden highlights opportunities to reduce health system costs and improve patient and family education.

We worked with parents of children with asthma to develop and evaluate two digital knowledge translation (KT) tools on asthma. These tools merge the best available research evidence with narratives of parent experiences, and use art and engaging media (video and interactive infographic) to optimize uptake and appeal. Following prototype completion, usability testing was conducted among 60 parents (30 parents per tool) in an urban Alberta emergency department waiting room. Parents viewed the tools on an iPad and answered questions via an electronic survey. Usability was assessed based on nine items with responses on a five-point Likert scale from 1=strongly disagree to 5=strongly agree. Overall, results were positive and the tools were highly rated across most usability items. Mean scores across usability items were 4.13 to 4.63 for the video and 4.10 to 4.43 for the infographic. The scores from the usability testing suggest arts-based digital tools are useful in sharing complex health information with parents about the care of a child with asthma and provide meaningful guidance on how to improve KT tools to better reflect the needs of parents of children with asthma.

## Introduction

Asthma is the most common chronic condition in children with an estimated 15% of children and youth living with asthma in Canada (1). The majority of asthma management occurs at home (2, 3), however management in children remains suboptimal (4-6) and poor control of symptoms has been reported in up to 75% of cases (7). Acute asthma exacerbations, or asthma attacks, are the main reason for children to seek emergency care, contributing to financial burdens for families and healthcare systems. This burden highlights opportunities to reduce health system costs and improve patient and family education.

A wide variety of asthma information is available including written and online materials. However, parents are not always aware of these resources, and even if they are aware, uncertainty about the credibility and reliability of information sources is high (8, 9). Additionally, resources provided often contain medical jargon as opposed to simplified lay language that is both understandable and useable for parents (10). Parental uncertainty combined with anxiety during a child’s illness are common, leading parents to seek emergency or medical care (8).

In a previously conducted mixed studies systematic review, we identified that parents lacked confidence in recognizing, treating, or seeking care for asthma exacerbations (11). Similarly, in a previously conducted qualitative descriptive study (publication in progress) we found that parents reported feelings of uncertainty as to when their child needed emergency care, and had a lack of basic information about how their children’s asthma medications worked and when to take them. Understanding these information needs is essential to creating resources that can effectively communicate health information to help parents make informed decisions when seeking health care for their asthmatic children. Furthermore, this information demonstrates that more effective knowledge translation (KT) tools are needed to satisfy parent information needs.

Previous research has demonstrated the positive impact of working with end-users of health information, such as parents and other caregivers, to develop KT tools (12-18). Such collaborations have resulted in tools that are relevant and meet the information needs of the appropriate stakeholders. More specifically, arts- and narrative-based KT tools have been proven to be effective sources of communication, translating complex health information into engaging and understandable content for parents (12-18). Therefore, we worked with parents of children with asthma to develop and evaluate two digital KT tools on asthma (video and interactive infographic) that were relevant to their information needs. Following prototype completion, usability testing was conducted using iPads in an urban Alberta emergency department (ED) waiting room. Final tools were made publicly available on our website (https://www.echokt.ca/tools) and disseminated through stakeholder websites and on social media.

## Methods

A series of studies using multiple methods and involving parent engagement were undertaken to develop, refine, and evaluate an animation video and interactive infographic for pediatric asthma. Research ethics approval was obtained from the University of Alberta Health Research Ethics Board (Edmonton, AB) [Pro00062904]. Operational approvals were obtained from the Stollery Children’s Hospital to conduct usability testing.

### Compilation of Parents’ Narratives

We conducted a qualitative descriptive study(19-21) involving semi-structured interviews (**Appendix A**) to identify parents’ experiences and information needs and to inform parental narratives for our tools. Parents with children who presented to the Stollery Children’s Hospital Emergency Department with asthma were recruited for qualitative interviews. Parents were asked to share their experiences with having a child with asthma. The qualitative descriptive study is currently being written for publication.

Concurrently, our research group conducted a mixed studies systematic review to synthesize current evidence about experiences and information needs of parents managing asthma. Detailed methods and results from the systematic review are published elsewhere (11).

### Prototype (Intervention) Development

Results from both the systematic review and qualitative interviews were used to inform the development of an infographic skeleton and video script. Clinical content from Bottom Line Recommendations (BLR) developed by TRanslating Emergency Knowledge for Kids (TREKK) was also included in the tools (22). Following the completion of the infographic skeleton and video script, researchers worked with illustrators and graphic designers to develop the tool prototypes.

### Video

The English-language video was 6 minutes and 35 seconds long, narrated in the third person, and included closed captioning. It outlined the story of a young child named Ari who has asthma. The video highlighted the process Ari took to become diagnosed with the condition and described important information about asthma, including symptoms, how it can be diagnosed, and how asthma may impact children differently. The video also outlined how asthma may be exacerbated by other conditions or triggers, such as viral infections, animals, dust mites, and smoke. Included in the video was information on asthma medications as well as when to seek emergency care or visit a doctor. Screen captures of the video are included in **Appendix B**.

### Infographic

The interactive infographic was developed in the same format as other infographics in our suite of tools (https://www.echokt.ca/tools/). The style is unique to our research program and was developed over the course of several years (23). The interactive infographic looks similar to a webpage and allows users to scroll through the information, exploring it at their own pace. The ability for parents to control what they view based on their needs differentiates the interactive infographic from the video. The information provided in the infographic mirrors the information provided in the video and is comprised of 8 major sections: (1) General Information about Asthma, (2) Diagnosis, (3) Symptoms, (4) Asthma Attacks, (5) When to seek Emergency Care vs When to go to a Doctor, (6) Treatment, (7) Triggers, (8) Managing Asthma Attacks. Within *Treatment*, the tool provides information about reliever medications, controller medications, and action plans. The infographic also provides useful links to other websites with information about asthma in children. Screen captures of the infographic are included in **Appendix C**.

### Revisions

Iterative processes were used to develop the tools and parents, health care professionals (HCPs), and researchers provided several rounds of feedback. HCPs were asked to comment on the accuracy of clinical information and evidence. Parents from our Pediatric Parent Advisory Group (P-PAG) (23) were asked to provide feedback on the length, stylistic elements, and information not addressed in the tools. The P-PAG meets once a month and members are asked to participate in tool development several times a year. Likewise, research team meetings are held weekly to discuss the development of our tools.

### Surveys

Parents presenting with an ill child to a major pediatric emergency department (ED) in the Edmonton area were recruited to participate in an electronic, usability survey (**Appendix D**). Members of the study team approached parents in the ED to determine interest and study eligibility. Parents who agreed to participate in the study were provided with an iPad by the researcher and asked to complete a consent form. Video usability testing was conducted from November 4, 2020 to November 21, 2020 and infographic testing was conducted from January 4, 2022 to January 20, 2022. Study team members were available in the ED to provide technical assistance and answer questions as parents were completing the surveys. The usability survey included 9, 5-point Likert items that assessed: 1) usefulness, 2) aesthetics, 3) length, 4) relevance, and 5) future use. The usability survey was designed in-house based upon key elements identified by a systematic search of over 180 usability evaluations (24). Parents were also asked to provide their positive and negative opinions of the tool via two free text boxes.

### Data Analysis

Data was cleaned and analyzed using SPSS v.24. Descriptive statistics and measures of central tendency were generated for demographic questions. Likert responses were given a corresponding numerical score, with 5 being “Strongly Agree” and 1 being “Strongly Disagree” (25, 26). Means and standard deviations (SDs) were calculated for each usability item. Independent two tailed t-tests were used to determine if there was a significant difference in the mean usability scores of the two KT tools for each usability item. Open-ended survey data was analyzed thematically.

Data was cleaned and analyzed using SPSS v.24. Descriptive statistics and measures of central tendency were generated for demographic questions. Likert answers were given a corresponding numerical score, with 5 being “Strongly Agree” and 1 being “Strongly Disagree”. Open-ended survey data was analyzed thematically.

See **Appendix E** for an overview of the entire project timeline.

## Results

Sixty parents awaiting pediatric ED care completed the usability survey (30 parents participated in the infographic usability testing and 30 parents participated in the video usability testing). The demographic characteristics of the parents who participated are presented in **Table 1**.

**Table 1.** Demographic characteristics of participants who assessed the usability of the asthma digital tools (video n=30; infographic n=30; combined n=60)

| Characteristic | Video<br>n (%) | Infographics<br>n (%) | Combined<br>n (%) |
| --- | --- | --- | --- |
| Sex |  |  |  |
| Female | 24 (80) | 24(80) | 48 (80) |
| Male | 6 (20) | 6(20) | 12 (20) |
| Ethnicity |  |  |  |
| African American/Canadian | 0 | 1 (3.3) | 1 (1.7) |
| Asian | 4 (13.3) | 3 (10) | 7 (11.7) |
| Black | 0 | 1 (3.3) | 1 (1.7) |
| East Indian | 0 | 1 (3.3) | 1 (1.7) |
| First Nations | 3 (10) | 0 | 3 (5) |
| Hispanic/Latino | 0 | 1 (3.3) | 1 (1.7) |
| Metis | 2 (6.7) | 0 | 2 (3.3) |
| Middle eastern/North African | 2 (6.7) | 3 (10) | 5 (8.3) |
| South Asian | 0 | 1 (3.3) | 3 (5) |
| White/Caucasian | 18 (60) | 17 (56.7) | 35 (58.3) |
| Prefer not to answer | 1 (3.3) | 0 | 1 (1.7) |
| Age |  |  |  |
| Less than 20 | 1 (3.3) | 0 | 1 (1.7) |
| 20-30 years | 1 (3.3) | 3 (10) | 4 (6.7) |
| 31-40 years | 18 (60) | 17 (56.7) | 35 (58.3) |
| 41-50 years | 7 (23.3) | 10 (33.3) | 17 (28.3) |
| 51-60 years | 2 (6.7) | 0 | 2 (3.3) |
| Missing | 1 (3.3) | 0 | 1 (1.7) |
| Marital Status |  |  |  |
| Married/Partnered | 24 (80) | 29 (96.7) | 53 (88) |
| Single | 6 (20) | 1 (3.3) | 7 (12) |
| Education |  |  |  |
| Some high school | 2 (6.7) | 0 | 2 (3.3) |
| High school diploma | 5 (16.7) | 4 (13.3) | 9 (15) |
| Some post-secondary | 3(10) | 3 (10) | 6 (10) |
| Post-secondary certificate/diploma | 14 (46.7) | 11(36.7) | 25 (42) |
| Post-secondary degree | 1 (3.3) | 7 (23.3) | 8 (13) |
| Graduate degree | 4 (13.3) | 5 (16.7) | 9 (15) |
| Other | 1 (3.3) | 0 | 1 (1.7) |
| Household Income |  |  |  |
| Less than \$25,000 | 2 (6.7) | 0 | 2 (3.3) |
| \$25,000-\$49,000 | 7 (23.3) | 2 (6.7) | 9 (15) |
| \$50,000-\$74,000 | 5 (16.7) | 3 (10) | 8 (13.3) |
| \$75,000-\$99,000 | 2 (6.7) | 7 (23.3) | 9 (15) |
| \$100,000-\$149,000 | 3(10) | 5 (16.7) | 8 (13.3) |
| \$150,000 and over | 4 (13.3) | 9 (30) | 13 (21.7) |
| Prefer not to answer | 7 (23.3) | 3 (10) | 10 (16.7) |
| Missing | 0 | 1 (3.3) | 1 (1.7) |
| Geographical Area |  |  |  |
| City | 26 (86.7) | 18 (60) | 44 (73.3) |
| Farm | 0 | 4 (13.3) | 4 (6.6) |
| Town | 0 | 1 (3.3) | 1 (1.7) |
| Suburb | 4 (13.3) | 7 (23.3) | 11 (18.3) |
| Number of Children in the Family |  |  |  |
| 0 | 0 | 1 (3.3) | 1 (1.7) |
| 1 | 10 (33.3) | 9 (30) | 19 (31.7) |
| 2 | 6 (20) | 13 (43.3) | 19 (31.7) |
| 3 | 11 (36.7) | 3 (10) | 14 (23.3) |
| 4 | 2 (6.7) | 4 (13.3) | 6 (10) |
| Missing | 1 (3.3) | 0 | 1 (1.7) |
| Relationship to child brought to the ED |  |  |  |
| Parent | 27(90) | 27(90) | 54 (90) |
| Guardian | 1 (3.3) | 1 (3.3) | 2 (3.3) |
| Other family member | 1 (3.3) | 1 (3.3) | 2 (3.3) |
| Missing | 1 (3.3) | 1 (3.3) | 2 (3.3) |
| Times Visited ED |  |  |  |
| 1-5 Times | 20(66.7) | 25(83.3) | 45 (75) |
| 6+ Times | 10 (33.3) | 4 (13.3) | 14 (23.3) |
| Missing | 0 | 1 (3.3) | 1 (1.7) |

Overall, parents scored the two tools positively with most selecting strongly agree and agree for the usability items (**Table 2**). The mean scores across usability items for the video ranged from 4.13 to 4.63 (on a 5-point scale with 4 indicating agree and 5 indicating strongly agree). Parents felt the video was useful and relevant to them. They found the video easy to use and felt it could be used without written instruction. Parents also found the length of the video appropriate and thought it was aesthetically pleasing. Parents agreed and strongly agreed that they would use the video in the future and that it would help them make decisions about their child’s health. Finally, when asked if they would recommend the video to a friend, parents agreed or strongly agreed.

**Table 2.** Means (SD) of participant responses to the usability survey

| Usability Measures | Video | Infographic | Significance (P-value) |
| --- | --- | --- | --- |
| It is useful. | 4.63 (0.49) | 4.37 (0.72) | 0.098 |
| It provides information that is relevant to me as a parent. | 4.57 (0.57) | 4.33 (0.66) | 0.148 |
| It is simple to use. | 4.60 (0.50) | 4.43 (0.50) | 0.203 |
| I can use it without written instructions or additional help. | 4.57 (0.50) | 4.43 (0.57) | 0.340 |
| Its length is appropriate. | 4.13 (0.86) | 4.27 (0.52) | 0.471 |
| It is aesthetically pleasing (i.e. images, colours, etc.). | 4.43 (0.63) | 4.40 (0.58) | 0.829 |
| It helps me make decisions about my child's health. | 4.40 (0.67) | 4.17 (0.65) | 0.177 |
| I would use it in the future. | 4.23 (0.77) | 4.10 (0.66) | 0.476 |
| I would recommend it to a friend. | 4.53 (0.51) | 4.17 (0.75) | 0.030* |

\*p value of <0.05 is considered significant.

For the infographic, mean scores across usability items ranged from 4.10 to 4.43 (on a 5-point scale with 4 indicating agree and 5 indicating strongly agree). Parents felt the infographic was useful and relevant to them. They found the infographic easy to use and felt it could be used without written instruction. Parents also found the length of the infographic appropriate and thought it was aesthetically pleasing. Parents agreed and strongly agreed that they would use the infographic in the future and that it would help them make decisions about their child’s health. Finally, when asked if they would recommend the infographic to a friend, parents agreed or strongly agreed.

There was a statistically significant difference in the mean ratings of the video versus the infographic on one of the usability items. Parents were more likely to recommend the video than the infographic to friends. There was no statistically significant difference for mean scores for the video versus infographic in terms of their usefulness, if they provided relevant information, if they were simple to use, if they could use them without written instructions or additional help, if their length was appropriate, if they were aesthetically pleasing, it they could help them make decisions about their child’s health, or if they should use them in the future.

There were a few negative comments in the open text responses about the video, which all centered around parents feeling the video was a bit long, however several parents also noted that “all the information was necessary”. In the positive comments, parents noted that the video was “very informative” and “easy to understand”. There were many positive comments where parents described the infographic as a “great tool” that was “very easy to read” and “easy to follow”. There were no negative comments about the infographic. As a result of these comments and the high rating of both tools, no revisions were made to the tools following usability testing.

## Conclusions

The purpose of this project was to develop two arts-based digital tools (an animation video and interactive infographic) for parents of children who have asthma and experience asthma exacerbations. Using a multi-method approach that encompassed stakeholder engagement, we were able to create tools that were highly rated amongst parents seeking care for their children in an urban emergency care centre. Parents found the tools to be useful, relevant, easy to use, and aesthetically pleasing. Most importantly, parents felt that the tools could facilitate decision-making in the future, with parents mainly agreeing and strongly agreeing that they would use the tool and recommend the tool to their friends. Results from this project indicate that end-user engagement plays a positive role in developing KT tools that address the needs of parents.

**The tools can be found here: <u>echokt.ca/asthma</u>**

Note: Our KT tools are assessed for alignment with current, best-available evidence every two years. If recommendations have changed, appropriate modifications are made to our tools to ensure that they are up-to-date (27).

## Other Outputs from this Project

### Research Papers

Shulhan-Kilroy J, Elliott SA, Scott SD, Hartling L. parents’ self-reported experiences and information needs related to acute pediatric asthma exacerbations: A mixed studies systematic review. PEC Innovation. 2022; 1:100006. doi: 10.1016/j.pecinn.2021.100006

## Supporting information

Supplemental Table 1: Adapted Reporting Checklist for Multi-Method research

## Data Availability

All data produced in the present study are available upon reasonable request to the authors

https://echokt.ca/asthma

## Author Contributions

This study was conducted under the supervision of Dr. Shannon D. Scott (SDS) and Dr. Lisa Hartling (LH), principal investigators (PIs) for **translating Evidence in Child Health to enhance outcomes** (ECHO) Research and the **Alberta Research Centre for Health Evidence** (ARCHE), respectively. Both PIs designed the research study and obtained research funding through the Canadian Institutes of Health Research (CIHR).

SDS designed and supervised all aspects of tool development and evaluation.

LH designed and provided input on all aspects of tool development and evaluation.

All authors contributed to the writing of this technical report and provided substantial feedback.

## Acknowledgements

Hannah Brooks (HB) conducted and analyzed qualitative interviews with parents.

Given COVID-19 pandemic restrictions which prevented ECHO research staff from collecting the usability data, University of Alberta Pediatric Emergency Medicine research staff were contracted to collect usability data.

Kathy Reid (KR) analyzed usability data.

**This work was funded by:**

**Canadian Institutes of Health Research:**

- Scott, S.D. (co-PI) & Hartling, L. (co-PI), Ali, S., Currie, G., Dyson, M., Fernandes, R., Fleck, B., Freedman, S., Jabbour, M., Johnson, D., Junker, A., Klassen, T., Maynard, D., Newton, A., Plint, A., Richer, L., Robinson, J., Robson, K., Vandall-Walker, V. [all collaborators listed in alphabetical order]. (2016) Integrating evidence and parent engagement to optimize Children’s healthcare. CIHR Foundation Scheme ($2,500,000). July 2016-June 2023.

## Appendices

### Appendix A – Qualitative Interview Guide

Parents will be interviewed to understand their experience having a child with an asthma exacerbation. Semi-structured interviews will be conducted with parents in order to get their “narrative” or experiences. The following questions will be used to guide these interviews. Being true to semi-structured interview techniques, interview questions will start broad and then move to the more specific.

1. Tell me about your experience having a child with asthma
  a. Tell me about the events <u>leading up</u> to your child being diagnosed. [Typically, the events leading up to being diagnosed take a good chunk of time]
    i. Signs/symptoms your child was experiencing (child’s health)
    ii. Visits to healthcare professionals to understand what was happening. (Probe number of visits, types of healthcare settings they visited, what did they do? Medications, assessments; were other diagnoses given to your child? E.g. allergies?)
    iii. Things you tried to decrease the symptoms (e.g., removing cats, removing carpets, no stuffed animals, etc.).
    iv. How did your <u>child feel</u> during this time? (emotions, mental state)
    v. How did <u>you feel</u> during this time? (emotions, mental state)
2. Tell me <u>when your child was diagnosed</u> with asthma.
  a. Confirm child’s age when diagnosed
  b. Who diagnosed your child with asthma?
  c. How did they diagnosis your child with asthma? (tests they did)
  d. What happened after they were diagnosed with asthma?
    i. Medications ordered? (tell me about the medications they take and response to medications)
    ii. Education given for day-to-day management
    iii. Follow up care provided?
    iv. Were there changes that the family needed to make for the child with asthma? (Probe – removing carpet from the house, removing cats from the house, quitting smoking, etc.)
3. Tell me about the <u>day-to-day management of your child’s asthma.</u>
  a. Probe for a sense of is the child’s asthma well managed or are their frequent flare ups.
  b. How do you feel about managing your child’s asthma (day to-day) – Probe for parent ‘s level of comfort/knowledge/skill being able to manage their child’s asthma
4. When your child’s asthma “flares up” (asthma exacerbation) – tell me how you manage it. (Probe if their child has had an exacerbation – not all children do)
  a. How do you know when your child is having a “flare up?” (probe child’s symptoms, signs)
  b. What do you do? (medications, assessments, seeking advice)
  c. When do you seek healthcare? (How do they decide to go for help? How do they decide where to go – medicentre or ED)?
  d. Do you feel that you have adequate knowledge, know-how to manage your child’s asthma flare ups?
  e. If you went to the ED, tell me about that experience (if there are many – suggest the last experience, or the most memorable experience).
    i. What assessments were done?
    ii. Medications
    iii. How was your child during the experience (emotions)?
    iv. How were you during the experience?
  f. Have the asthma “flare ups” affected other aspects of your child’s life?
    i. School attendance
    ii. Friends/Social Support
5. Are there “things” that make it challenging to care for your child with asthma?
  a. Probe for – knowledge needs? (If yes, what would they like to know more about); is it parental knowledge needs or the need for others in their child’s live to know more about asthma (e.g., teachers, coaches, grandparents).
  b. Environmental challenges (where they live, where the child goes to school, weather/seasonal challenges)
  c. Personal challenges (the costs associated with having a child with asthma)

**Appendix B.**
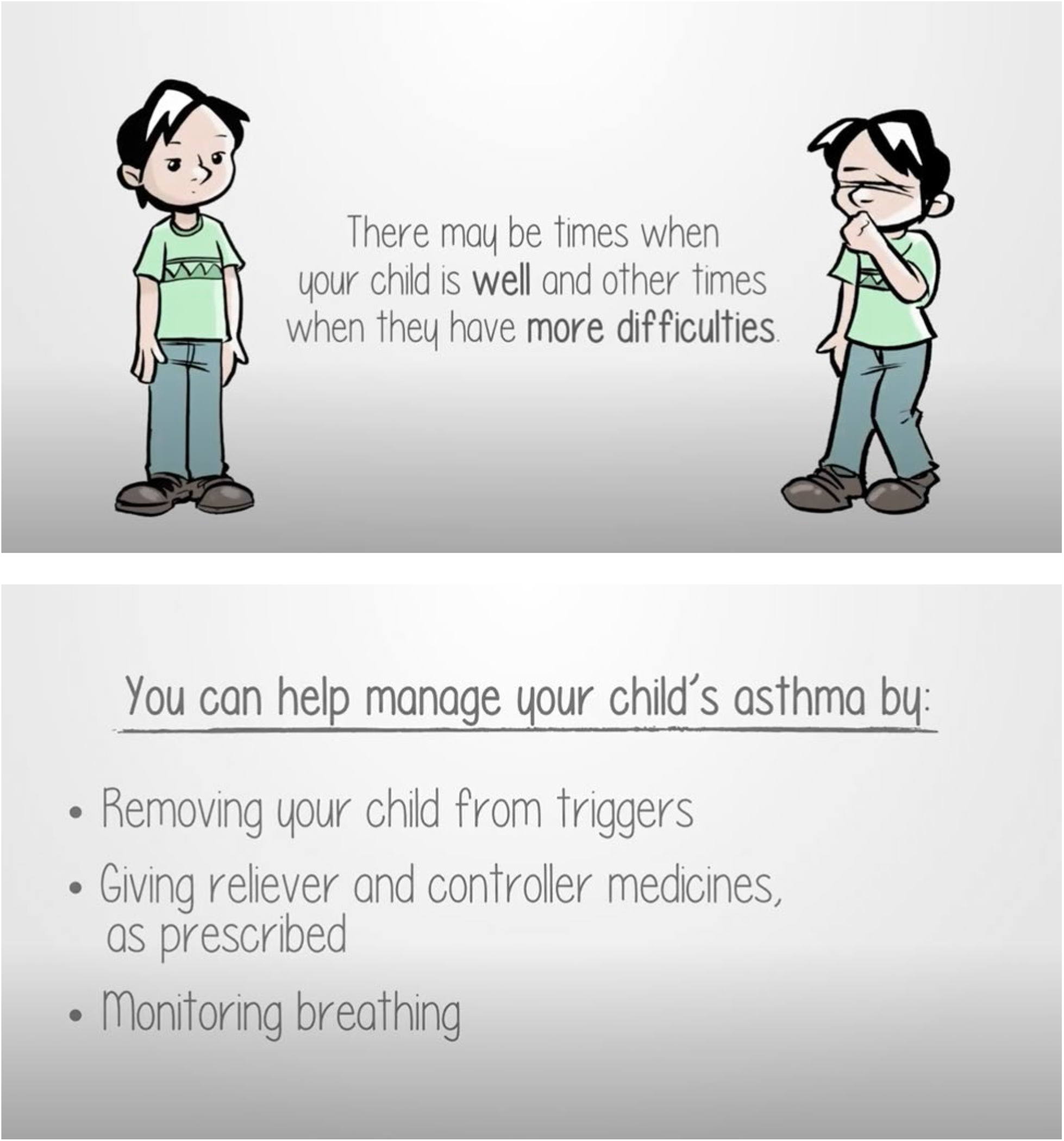
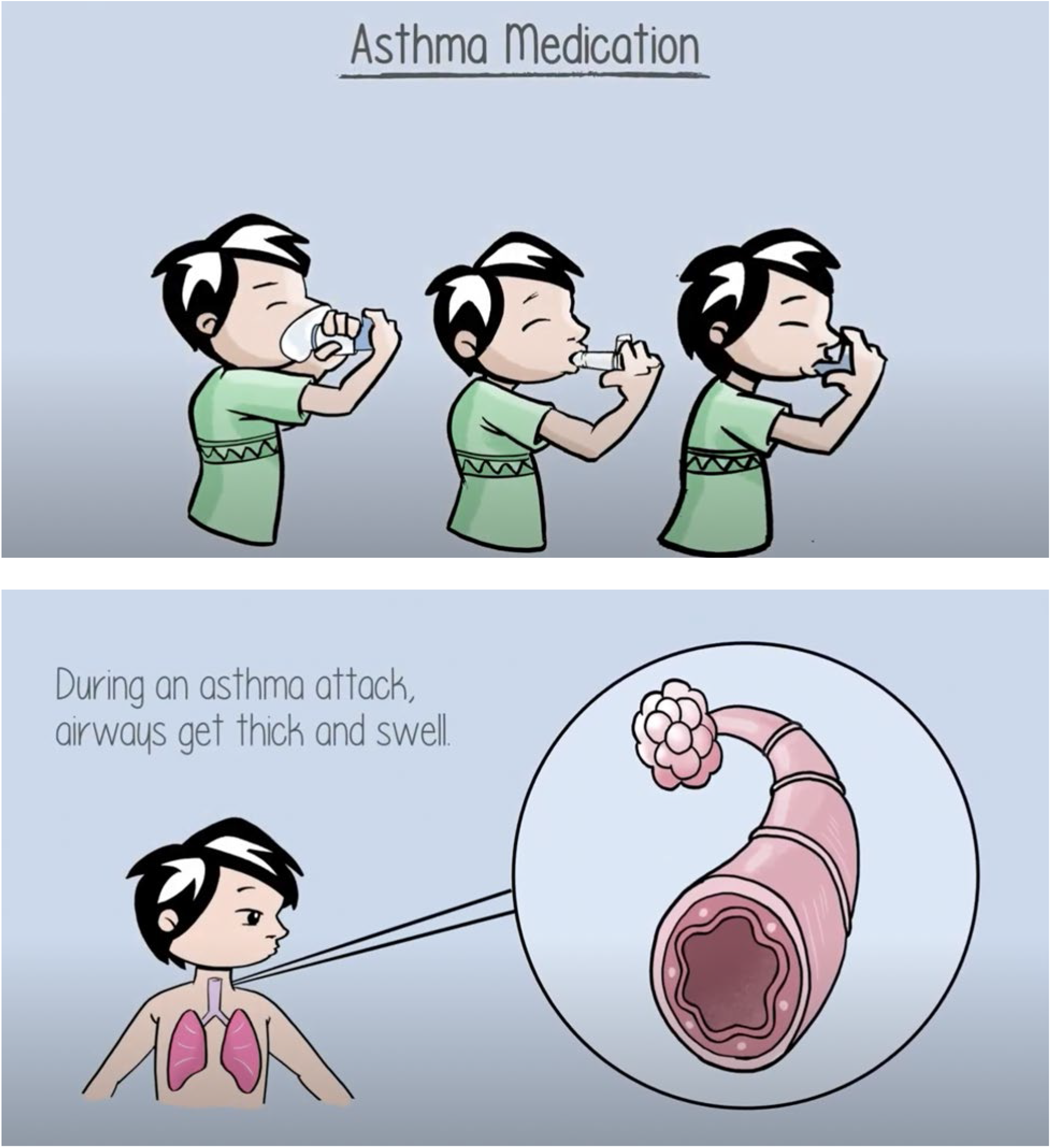
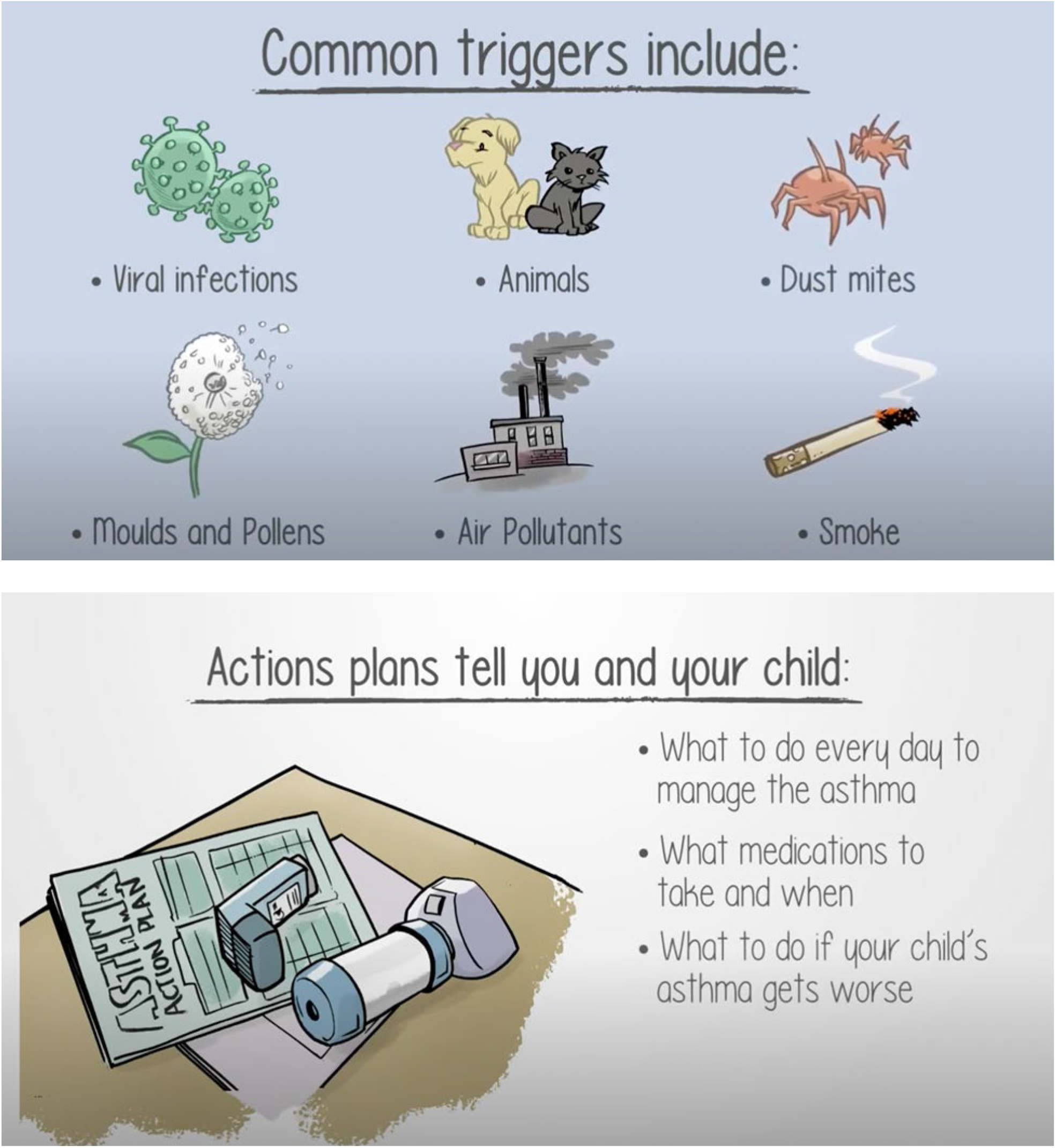
Images from art-based video about childhood asthma (video available at https://www.echokt.ca/asthma/)

**Appendix C.**
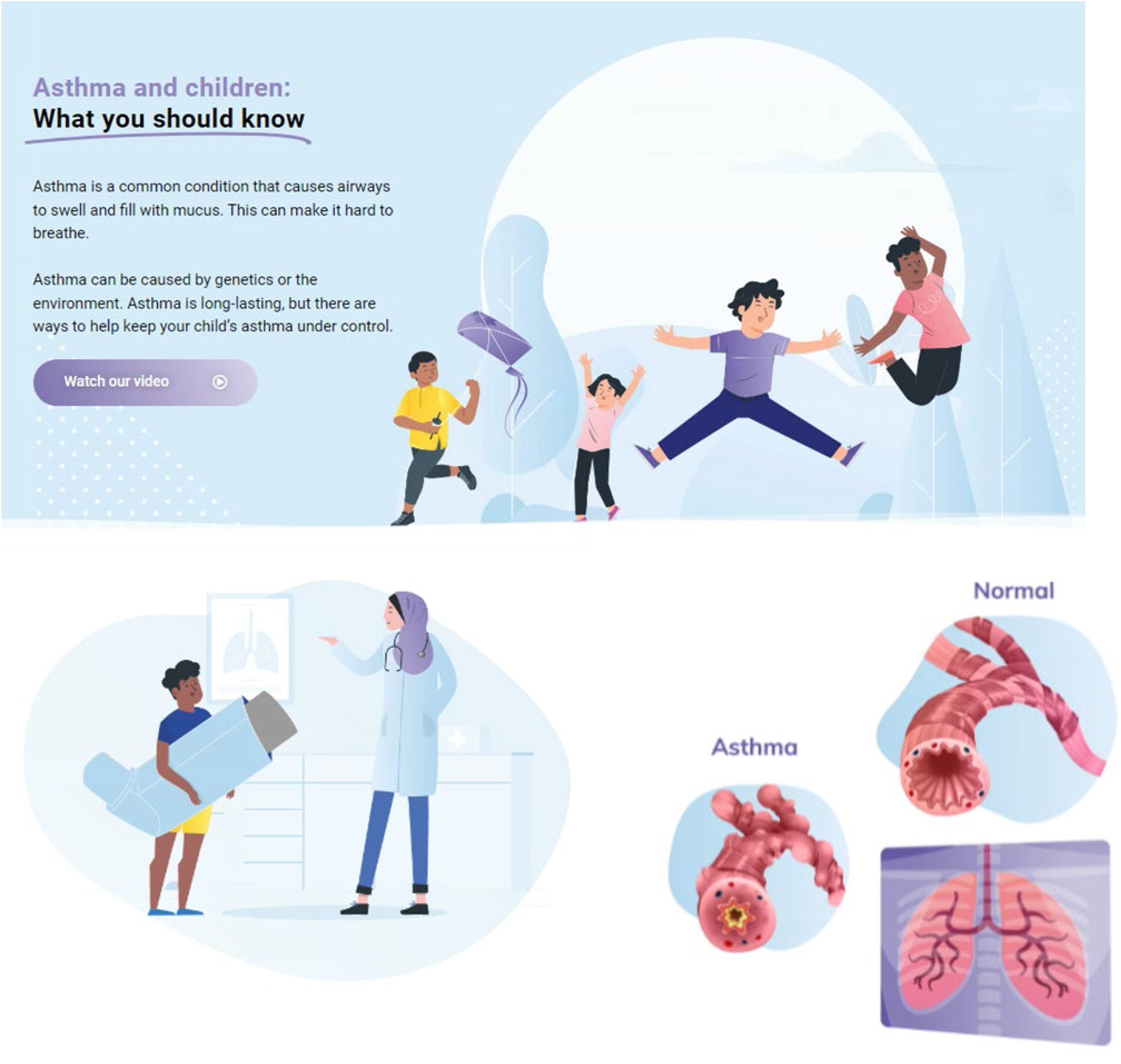
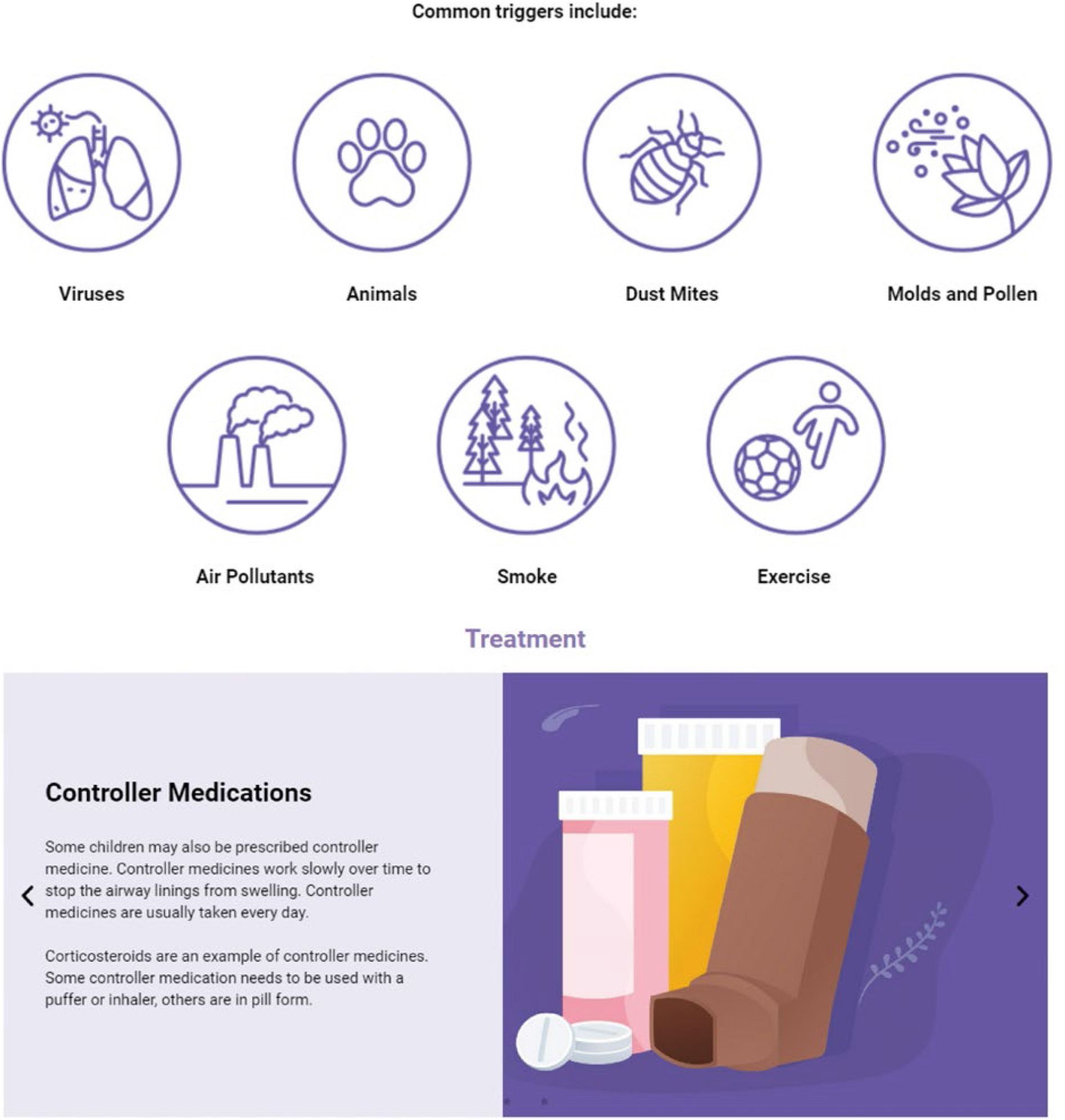
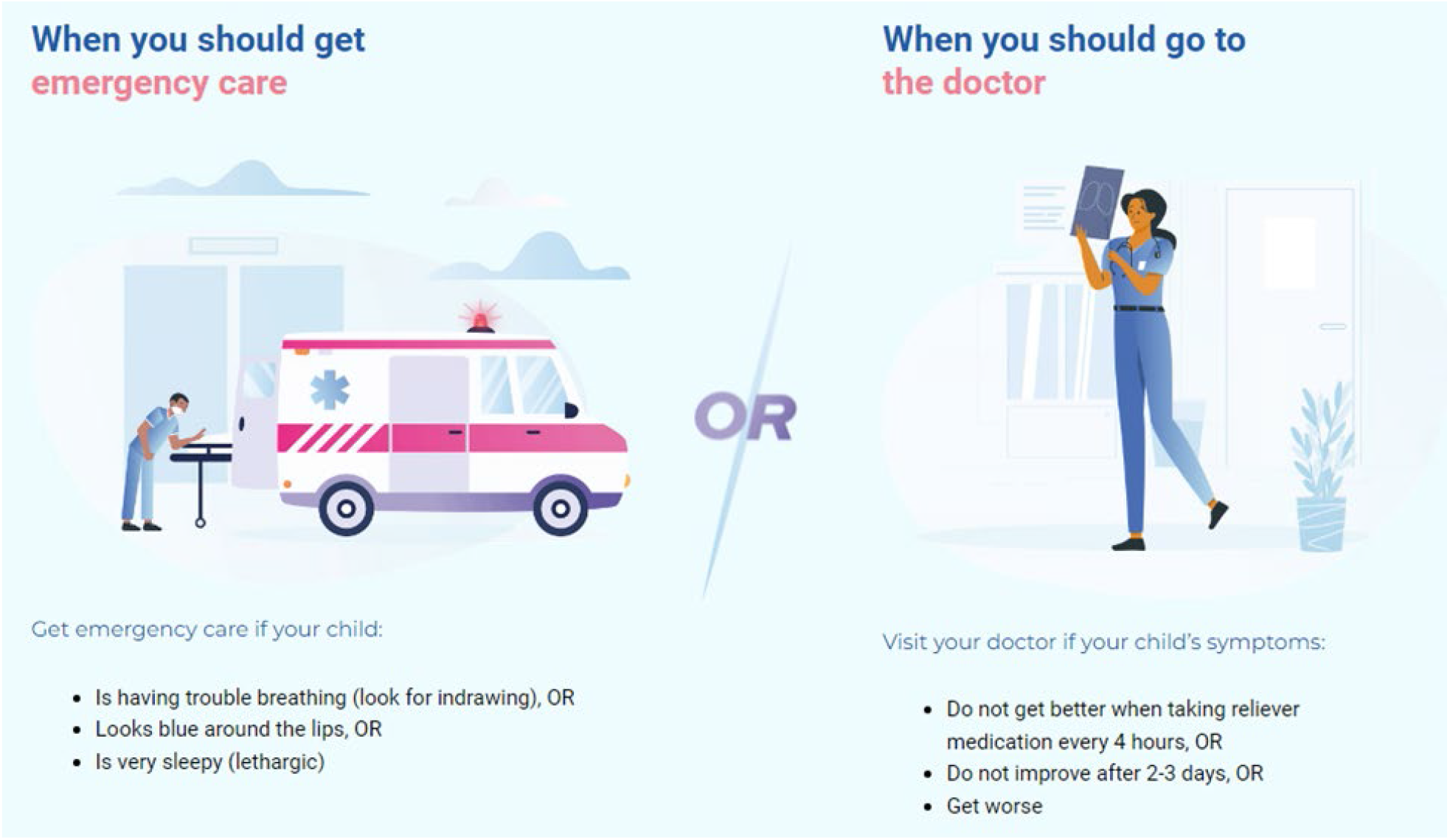
Images from Infographic about childhood asthma (infographic available at https://www.echokt.ca/asthma-infographic/)

### Appendix D – Usability Survey

SECTION 1: Demographics

1) a. What is your gender?
  □ Male
  □ Female
  □ Non-binary
  □ Two-spirit
  □ Other:______
  □ Prefer not to answer
1) b. Would you describe yourself as transgender?
  □ Yes
  □ No
  □ Prefer not to answer
2) Which ethnicities best describes you? *Please select all that apply*.
  □ Asian
  □ African American or African Canadian
  □ Black
  □ First Nations
  □ Hispanic or Latino
  □ Métis
  □ Middle Eastern or North African
  □ South Asian
  □ Southeast Asian
  □ White or Caucasian
  □ Not listed:______
  □ Prefer not to answer
3) What is your Age?
  □ Less than 20 years old
  □ 20-30 years
  □ 31-40 years
  □ 41-50 years
  □ 51 years and older
4) What is your Marital Status?
  □ Married/Partnered
  □ Single
5) What is your gross annual household income?
  □ Less than $25,000
  □ $25,000-$49,999
  □ $50,000-$74,999
  □ $75,000-$99,999
  □ $100,000-$149,999
  □ $150,000 and over
  □ Prefer not to answer
6) What is your highest level of education?
  □ Some high school
  □ High school diploma
  □ Some post-secondary
  □ Post-secondary certificate/diploma
  □ Post-secondary degree
  □ Graduate degree
  □ Other:______
7) Where does your household live
  □ City
  □ Suburb
  □ Town
  □ Farm
  □ Other:______
8) What is your relationship to the child that you have brought to the emergency department?
  □ Parent
  □ Grandparent
  □ Other family member
  □ Guardian
9) How many children do you have? ______
10) How old are your children? ______
11) How many times have you visited the emergency department with your children?
  □ 1-5 times
  □ 6+times
12) Have any of your children ever been admitted to the hospital?
  □ Yes
  □ >No

SECTION 2: Assessment of attributes of the arts-based, digital tools

Note: items 1-9 are rated on a 5-point Likert scale from 1=strongly disagree to 5=strongly agree

1. It is useful. [5-point Likert Scale]
2. It provides information that is relevant to me as a parent. [5-point Likert Scale]
3. It is simple to use. [5-point Likert Scale]
4. I can use it without written instructions or additional help. [5-point Likert Scale]
5. Its length is appropriate. [5-point Likert Scale]
6. It is aesthetically pleasing (i.e., images, colours, etc.). [5-point Likert Scale]
7. It helps me to make decisions about my child’s health. [5-point Likert Scale]
8. I would use it in the future. [5-point Likert Scale]
9. I would recommend it to a friend. [5-point Likert Scale]
10. List the most negative aspects: [open text]
11. List the most positive aspects: [open text]

**Appendix D.**
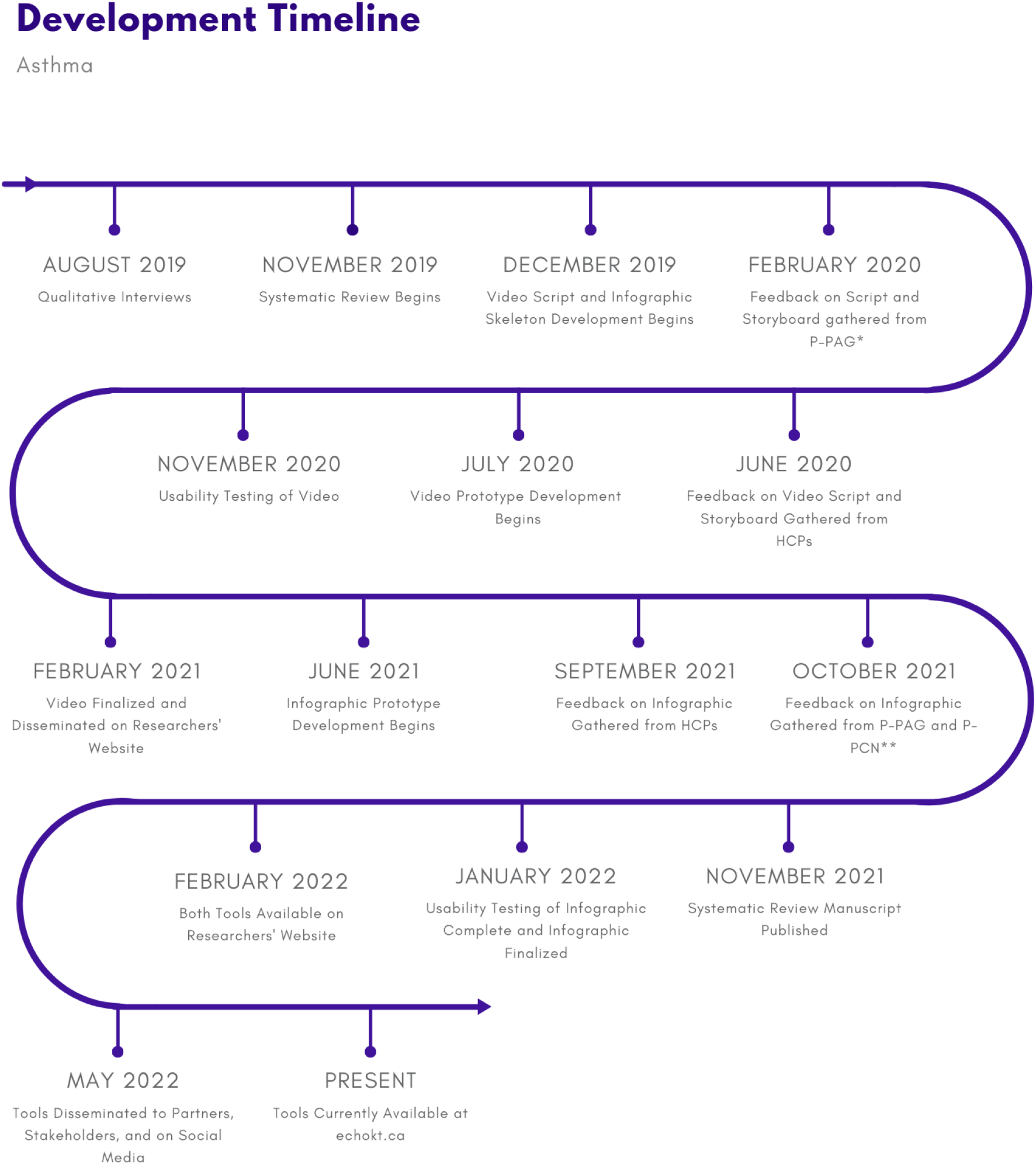
Project Timeline.

*P-PAG = Parents from our Pediatric parents’ Advisory Group (P-PAG)

**P-PCN = Pediatric Parent Consultation Network

HCPs = Healthcare Providers

## References

1. Canadian Institute for Health Informatin (CIFHI). Asthma Hospitalizations Among Children and Youth in Canada: Trends and Inequalities [Internet]. Ottawa, ON.[cited 2023 Feb 13]. Available from: https://www.cihi.ca/sites/default/files/document/asthma-hospitalization-children-2018-chartbook-en-web.pdf

2. Nicholas DB, Dell SD, Fleming-Carroll B, Selkirk EK. An evaluation of pediatric asthma educational resources. Social Work in Health Care. 2009;48(4):450–61. doi: 10.1080/00981380802589936

3. Deis JN, Spiro DM, Jenkins CA, Buckles TL, Arnold DH. Parental knowledge and use of preventive asthma care measures in two pediatric emergency departments. Journal of Asthma. 2010;47(5):551–6. doi: 10.3109/02770900903560225

4. Peterson-Sweeney K, McMullen A, Yoos HL, Kitzmann H, Halterman JS, Arcoleo KS, et al. Impact of asthma education received from health care providers on parental illness representation in childhood asthma. Research in Nursing and Health. 2007;30(2):203–12. doi: 10.1002/nur.20182

5. Stewart M, Anderson S, McGhan S, Letourneau N, Masuda JR. Online solutions to support needs and preferences of parents of children with asthma and allergies. Journal of Family Nursing. 2011;17(3):357-79-79. doi: 10.1177/1074840711415416

6. Swerczek LM, Banister C, Bloomberg GR, Bruns JM, Epstein J, Highstein GR, et al. A Telephone Coaching Intervention To Improve Asthma Self-Management Behaviors. Pediatric Nursing. 2013;39(3):125–45.

7. Mcghan SL, MacDonald C, James DE, Naidu P, Wong E, Sharpe H, et al. Factors Associated with Poor Asthma Control in Children Aged Five to 13 years. Canadian Respiratory Journal. 2006;13(1):23–9. doi: 10.1155/2006/149863

8. Neill SJ. Acute childhood illness at home: The parents’ perspective. Journal of Advanced Nursing. 2000;31(4):821–32. doi: 10.1046/j.1365-2648.2000.01340.x

9. Jackson C, Cheater FM, Reid I. A systematic review of decision support needs of parents making child health decisions. Health Expectations. 2008;11(3):232–51. doi: 10.1111/j.1369-7625.2008.00496.x

10. Samuels-Kalow M, Hardy E, Mollen C, Rhodes K, Uspal J, Reyes Smith A. Unmet Needs at the Time of Emergency Department Discharge. Academic Emergency Medicine. 2016;23(3):279–87. doi: 10.1111/acem.12877

11. Shulhan-Kilroy J, Elliott SA, Scott SD, Hartling L. parents’ self-reported experiences and information needs related to acute pediatric asthma exacerbations: A mixed studies systematic review. PEC Innovation. 2022;1:100006. doi: 10.1016/j.pecinn.2021.100006

12. Archibald MM, Scott SD. Learning from usability testing of an arts -based knowledge translation tool for parents of a child with asthma. Nursing Open. 2019;6(4):1615–25. doi: 10.1002/nop2.369

13. Meherali S, Hartling L, Scott SD. Cultural Adaptation of Digital Knowledge Translation Tools for Acute Otitis Media in Low- to Middle-Income Countries: Mixed Methods Usability Study. JMIR formative research. 2021;5(1):e13908. doi: 10.2196/13908

14. Reid K, Le A, Norris A, Scott SD, Hartling L, Ali S. Development and usability evaluation of an art and narrative-based knowledge translation tool for parents with a child with pediatric chronic pain: Multi-method study. Journal of Medical Internet Research. 2017;19(12). doi: 10.2196/jmir.8877

15. Scott SD, L. A, Hartling L. Developing and testing two arts-based knowledge translation tools for parents about pediatric acute gastroenteritis. Internal Technical Report. ECHO Research, University of Alberta. MedRxiv; 2021. doi: 10.1101/2021.06.08.21258514

16. Scott SD, L. A, Hartling L. Developing and testing an arts-based, digital knowledge translation tool for parents about childhood croup. Internal Technical Report. ECHO Research, University of Alberta. MedRxiv; 2021. doi: 10.1101/2021.06.03.21257424

17. Hartling L, Scott SD, Johnson DW, Bishop T, Klassen TP. A Randomized Controlled Trial of Storytelling as a Communication Tool. PLoS ONE. 2013;8(10). doi: 10.1371/journal.pone.0077800

18. Scott SD, O ‘Leary KA, Archibald M, Hartling L, Klassen TP. Stories - a novel approach to transfer complex health information to parents: A qualitative study. Arts and Health. 2012;4(2):162-73-73. doi: 10.1080/17533015.2012.656203

19. Sandelowski M. Focus on research methods: Whatever happened to qualitative description? Research in Nursing and Health. 2000;23(4):334–40. doi: 10.1002/1098-240x(200008)23:4<334::aid-nur9>3.0.co;2-g

20. Sandelowski M. What ‘s in a name? Qualitative description revisited. Research in Nursing and Health. 2010;33(1):77–84. doi: 10.1002/nur.20362

21. Bradshaw C, Atkinson S, Doody O. Employing a Qualitative Description Approach in Health Care Research. Global Qualitative Nursing Research. 2017;4. doi: 10.1177/2333393617742282

22. Translating Emergy Knowledge for Kids (TREKK). Bottom Line Recommendations: Asthma Version 1.5 [Internet].Winnipeg, Manitoba: TREKK; 2020 January [cited 2023 February 13]. Available from: https://trekk.ca/system/assets/assets/attachments/439/original/2020-01-23_Asthma_BLR_v_1.5.pdf?1579801436

23. Hartling L, Elliott SA, Buckreus K, Leung J, Scott SD. Development and evaluation of a parent advisory group to inform a research program for knowledge translation in child health. Research Involvement and Engagement. 2021;7(1). doi: 10.1186/s40900-021-00280-3

24. Hornbæk K. Current practice in measuring usability: Challenges to usability studies and research. International Journal of Human Computer Studies. 2006;64(2):79–102. doi: 10.1016/j.ijhcs.2005.06.002

25. Carifio J, Perla R. Resolving the 50-year debate around using and misusing Likert scales. Medical Education. 2008;42(12):1150–2. doi: 10.1111/j.1365-2923.2008.03172.x

26. Sullivan GM, Artino AR, Jr. Analyzing and interpreting data from likert-type scales. J Grad Med Educ. 2013;5(4):541–2. doi: 10.4300/JGME-5-4-18

27. Featherstone RM, Leggett C, Knisley L, Jabbour M, Klassen TP, Scott SD, et al. Creation of an Integrated Knowledge Translation Process to Improve Pediatric Emergency Care in Canada. Health Communication. 2018;33(8):980–7. doi: 10.1080/10410236.2017.1323538

