## Supplemental Table 1: Adapted Reporting Checklist for Multi-Method research for "The development and usability testing of two arts-based knowledge translation tools for pediatric asthma"

**Currently there is no reporting checklist suggested by the EQUATOR Network for Multi-Methods Studies. As a result, Lee et al. (2022) checklist for mixed methods research was adapted for the purpose of reporting this Multi-Method Study. The original checklist is displayed in Table 1 on the left and adaptations to the checklist are shown on the right of the table. The adaptations are highlighted in red. This study is reported using the adapted checklist on the right of the table. **

**Table 1: Adaptation of a Mixed Methods Research Checklist to Multimethod Research Checklist for the Purpose of Reporting**

| **Checklist for Mixed Method Research (MMR) Manuscript preparation and review** | | **Applicable or Not Applicable to Multi-Method Research** | **Adapted Checklist for Multimethod Research**  **Manuscript preparation and review** | |
| --- | --- | --- | --- | --- |
| Rationale and description MMR Design | ☐Provide a Clear statement of the study Purpose | Applicable | Rationale and description of multi-method design | ☒Provide a Clear statement of the study Purpose |
|  | ☐Explicitly describe the MMR design in accordance with Creswell’s typology and use a diaphragm to illustrate the relationship and sequence of qualitative and quantitative research components | N/A- change suggested |  | ☒Explicitly describe the multi-method design and use a diagram to illustrate the sequence of qualitative and quantitative research methods |
|  | ☐Justify why the MMR design is appropriate for meeting the study purpose | Applicable with slight change |  | ☒Justify why the Multi-method design is appropriate for meeting the study purpose |
| Transparency in describing method details | ☐Describe the study population(s) and sample (e.g., who, what, how many) | Applicable | Transparency in describing method details | ☒Describe the study population(s) and sample (e.g., who, what, how many) |
|  | ☐Describe the sampling procedures (including inclusion and exclusion criteria, recruitment) | Applicable |  | ☒Describe the sampling procedures (including inclusion and exclusion criteria, recruitment) |
|  | ☐Describe qualitative data collection processes (how often data were collected, who collected the data, what kind of data collection instruments were used, how data were recorded—e.g., notes, transcripts) | Applicable |  | ☒Describe qualitative data collection processes (how often data were collected, who collected the data, what kind of data collection instruments were used, how data were recorded—e.g., notes, transcripts).  Comment from authors: ** Full report to be available soon, manuscript being prepared for publication** |
|  | ☐Describe quantitative data collection processes (how often data were collected, who collected the data, what kind of data collection instruments were used measurements, validity/reliability) | Applicable |  | ☒Describe quantitative data collection processes (how often data were collected, who collected the data, what kind of data collection instruments were used measurements, validity/reliability |
|  | ☐Describe qualitative data analysis processes (coding, single or multiple coders, replication logic, credibility) | Applicable |  | ☒Describe qualitative data analysis processes (coding, single or multiple coders, replication logic, credibility)  Comment from authors: ** Full report to be available soon, manuscript being prepared for publication** |
|  | ☐Describe quantitative data analysis procedures (missing data and how they are handled, statistical tests used) | Applicable |  | ☒Describe quantitative data analysis procedures (missing data and how they are handled, statistical tests used). |
| Integration of qualitative and quantitative research component | ☐Interpret qualitative analysis results with appropriate quotes if necessary | Applicable | Explain how the data is analyzed at each stage. Explain how the one stage of the research informed the next stage of the research | ☒Interpret qualitative analysis results with appropriate quotes if necessary.  Comment from authors: ** Full report to be available soon, manuscript being prepared for publication** |
|  | ☐Interpret quantitative analysis results in consideration of statistical significance, selection bias, and threats to validity | Applicable |  | ☒Interpret quantitative analysis results in consideration of statistical significance, selection bias, and threats to validity |
|  | ☐Compare qualitative and quantitative results | Not applicable |  | ☒Explain how the one stage of the research informed the next stage of the research |
|  | ☐Address divergencies and inconsistencies between qualitative and quantitative results | Not applicable |  |  |

Adapted from: Lee SD, Iott B, Banaszak-Holl J, Shih SF, Raj M, Johnson KE, Kiessling K, Moore-Petinak N (2022). Application of Mixed Methods in Health Services Management Research: A Systematic Review. Med Care Res Rev. 2022;79(3):331-344. <https://doi.org/10.1177/10775587211030393>
